# Perceptions and compliance with COVID-19 preventive measures in Southern and Central regions of Mozambique: a quantitative in-person household survey in the districts of Manhiça and Quelimane

**DOI:** 10.1101/2022.11.17.22282473

**Authors:** Ariel Nhacolo, Amílcar Magaço, Felizarda Amosse, Aura Hunguana, Teodomiro Matsena, Arsénio Nhacolo, Elisio Xerinda, Quique Bassat, Charfudin Sacoor, Inácio Mandomando, Khátia Munguambe

**Affiliations:** Centro de Investigação em Saúde de Manhiça (CISM), Maputo, Mozambique; ISGlobal, Hospital Clínic - Universitat de Barcelona, Barcelona, Spain; ICREA, Pg. Lluís Companys 23, 08010 Barcelona, Spain; Pediatrics Department, Hospital Sant Joan de Déu, Universitat de Barcelona, Barcelona, Spain; Consorcio de Investigación Biomédica en Red de Epidemiología y Salud Pública (CIBERESP), Madrid, Spain; National Institute of Health, Ministry of Health, Maputo, Mozambique; Faculty of Medicine, Eduardo Mondlane University, Maputo, Mozambique

## Abstract

**Background:** The COVID-19 pandemic has led countries into urgent implementation of stringent preventive measures at the population level. However, implementing these measures in low-income countries like Mozambique was incredibly difficult, coupled with lack of scientific evidence on the community understanding and compliance with these measures. This study assessed the perceptions and implementation of COVID-19 preventive measures recommended by Mozambican authorities in Manhiça and Quelimane districts, taking confinement, social distancing, frequent handwashing, mask wearing, and quarantine as the key practices to evaluate.

**Methods:** A quantitative survey interviewing households’ heads in-person was conducted in October 2020 and February 2021; collecting data on perceptions of COVID-19, symptoms, means of transmission/prevention; including self-evaluation of compliance with the key measures, existence of handwashing facilities, and the ratio of face-masks per person. The analysis presents descriptive statistics on perceptions and compliance with anti-COVID-19 measures at individual and household levels, comparing by district and other variables. T-test was performed to assess the differences on proportions between the districts or categories of respondents in the same district.

**Results:** The study interviewed 770 individuals of which 62.3% were heads of households, 18.6% their spouses, and 11.0% sons/daughters. Most participants (98.7%) had heard of COVID-19 disease. The most difficult measure to comply with was staying at home (35.8% of respondents said they could not comply with it at all); followed by avoiding touching the month/nose/eyes (28.7%), and social distancing at home (27.3%). Mask wearing in public places was the measure that more respondents (48.8%) thought they complied 100% with it, followed by avoiding unnecessary traveling (40.0%), avoiding crowed places (34.0%), and social distancing outside home (29.0%). Only 30.4% of households had handwashing devices or disinfectant (36.7% in Manhiça and 24.1% in Quelimane); and of those with devices, only 41.0% had water in the device, 37.6% had soap, and 22.6% had other disinfectant. The ratio of masks per person was only 1, which suggests that people may have used the same mask for longer periods than recommended.

**Conclusions:** Community members in Manhiça and Quelimane were aware of COVID-19 but they lacked understanding for implementing the preventive measures. This, together with socio-economic constraints, led to lower levels of compliance with the key measures. Understanding and addressing the factors affecting proper implementation of these measures is crucial for informing decision-makers about ways to improve community knowledge and practices to prevent infectious diseases with epidemic potential.

## 1. INTRODUCTION

Since COVID-19 was declared a pandemic disease (1), different approaches to contain the spread of the virus have been adopted around the world, with countries taking stringent measures which included, among others, closing the borders, banning incoming flights, shutting down institutions and non-essential services, curfews, restriction on opening hours of major services, enforcing quarantines to the infected and potential contacts, as well as extreme measures such as complete population lock-down (2)(3). While the implementation of these measures in high-income countries was already challenging, in low-income countries like Mozambique they represented an incredible conundrum. Mozambique initially declared level 3 State of Emergency, which included measures such as closing down educational institutions, interrupting visa services, limiting mobility, limiting gatherings to a maximum of 50 people and recommending a minimum distance of 1.5 meters between individuals. Later on, gatherings were limited to a maximum of 10 people and the use of masks was recommended on crowed places and public transport vehicles (4). The Mozambican Government focused on prevention, recognising that the health care system was far from being capable of responding to a massive number of COVID-19 patients, should the pandemic reach its peak early and abruptly (4).

However, and even during the harshest restriction periods, while facing hard confinement measures, there were reports, on the media, of crowded public transports, markets and streets, and of people not having interrupted activities such as working, or trading or travelling for the sake of their families’ livelihood (5). For the same reasons of self and family survival, there were indications that the “stay at home” principle was challenging to an important segment of the population, who lives on the basis of a daily income (6). Little is known about whether and how the communities understood and implemented the measures recommended for preventing or reducing the spread of COVID-19 in Mozambique.

Understanding the gaps in knowledge and compliance with measures against the spread of COVID-19 in Southern and Central Mozambique would confer the opportunity to develop more effective and socio-culturally appropriate sensitization initiatives to address the need of COVID-19 prevention. This study aimed to assess the perceptions and implementation of the measures recommended by the government of Mozambique to prevent COVID-19 in rural and urban settings of Southern and Central Mozambique (Manhiça and Quelimane districts), taking confinement, social distancing, hand washing, mask wearing, and quarantine as the key practices to assess. Data such as these, from both rural and urban areas and from two socio-economically different region of the country, were crucial for informing decision-makers about ways to improve community knowledge and practices regarding prevention of COVID-19 or any other future infectious disease with epidemic potential.

## 2. METHODS

### 2.1. Study design

This study was a cross-sectional quantitative household survey designed to collect data both at individual and household level, through interviews administered in-person to the heads of households or their representatives, gathering their perceptions and practices regarding anti-COVID-19 measures and in the context of their households.

### 2.2. Study setting and population

The study took place in two locations: (i) Manhiça district, a mainly rural setting located in Province Maputo, in the Southern region of Mozambique; and (ii) Quelimane district, in the Central province of Zambézia, which comprises urban and rural settings (Figure 1). Manhiça district is 85 km North of Maputo City, the capital of Mozambique, and was purposively selected because of the presence of the Manhiça Health Research Center (CISM). CISM has been conducting biomedical research in the district over nearly 25 years, which has facilitated the implementation of the study in a context of emergency with relatively less challenges than it would have been elsewhere in rural Mozambique.

**Figure 1.**
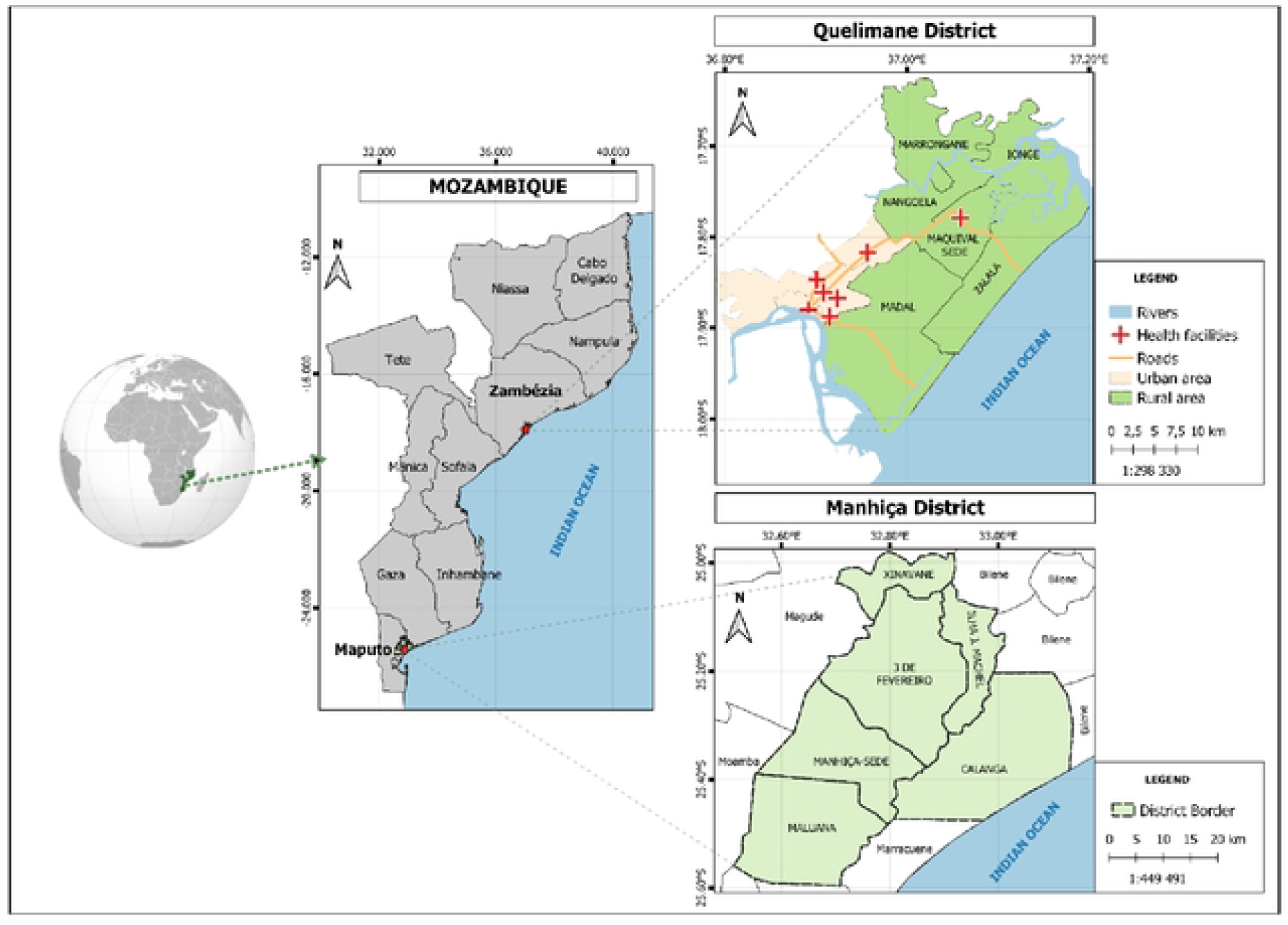
Location and administrative division of Manhiça and Quelimane districts. Source: Manhiça and Quelimane HDSS databases (2022) and (10).

Similarly, the district of Quelimane was chosen because CISM has been conducting biomedical research there for 5 years, particularly on causes of deaths using minimally invasive tissue sampling (7). Currently, CISM is establishing an HDSS in the district. Quelimane is located along the river Rio dos Bons Sinais in the Southern part of Zambézia province. The district has urban and rural settings. The urban area comprises the City of Quelimane (the capital of Zambézia province), where live 71.7% of the total 349,842 district population (8).

The study population comprised heads of households or their representatives that were residents in the study area, as defined by the CISM’s HDSS, i.e. those who live in a household in Manhiça or in Quelimane districts for three or more months or are entering the district with intention for that (9). The survey adopted, also, the HDSS definition of household, as a group of one or more individuals who live together in the same house or group of houses, eat together, share domestic expenses, and acknowledge one of them as their head or leader (9). The head of a household is the member who takes the most important day-to-day decisions in the household and is the reference member (9).

### 2.3. Sampling and sample size

In Manhiça, the survey used the HDSS database as a sampling frame to randomly select a sample of households that was representative at the level of Administrative Posts. In Quelimane the sampling frame was a list of households given by the local authorities. Although the study had multiple outcomes of interest at household level, the presence of a handwashing facility with water and soap was considered as the main variable for estimating the minimum sample size of households for this study. A handwashing facility was defined as a device to contain, transport or regulate the flow of water to facilitate handwashing (11) -a commonly used proxy indicator of actual handwashing practice, which has been found to be more accurate than other proxies such as self-reports of hand washing practices. Because there were no data on the proportion of households with a handwashing facility during the pandemic in Mozambique, the sample size was calculated to estimate a proportion of 50% with a margin of error of 5% and confidence level of 95% (12). Thus, it was estimated that a sample size of 385 households for each district would be sufficient to estimate this proportion (a sample size calculated this way is also suitable to estimate proportions ranging from 10% to 90%) (12). At the individual level, the main outcome was defined as the proportion of individuals who wash their hands at critical points in time, but because there were no data for the pandemic period, the sample size was calculated to estimate a proportion of 50% of people washing their hands, with a margin of error of 5% and confidence level of 95%, which resulted in a sample size of 385 individuals for each district. Thus, because the sample size for households coincided with that of individuals, the data for the two units of analysis (individual and household) were collected by asking the questions to the same respondent, i.e. one respondent per household.

### 2.4. Data collection and quality assurance

The data were collected in October 2020 in Manhiça and in February 2021 in Quelimane, using paper-based questionnaires that were verified by demographers for consistencies and completeness, and were double-entered to reduce typing errors at CISM’s Data Center. Forms with errors were returned to the field for corrections. The interviewers were selected from the HDSS fieldworkers (supervisors and other well-experienced fieldworkers) who were carefully trained for this survey. The training included refreshing the training that they had previously received on biosafety measures in the context of the coronavirus pandemic, as part of CISM’s requirements. The survey questionnaire was designed to collect the following data:

*<u>Perceptions and level of compliance with COVID-19 preventive measures</u>* – using questions framed to capture knowledge and understanding of the disease, details of symptoms, means of transmission and prevention, definitions of preventive measures, and level of compliance with anti-COVID-19 measures. Thus, questions such as “have you ever heard of coronavirus?”; “have you ever heard of COVID-19”; “if YES, what it is?”; “what are the symptoms or signs of Coronavirus or COVID-19?”; “how is this disease transmitted?”; and “how can we prevent it?” were asked. To assess the understanding of anti-COVID-19 measures, respondents were asked the following questions: “could you please explain what does social distancing means?”; “could you explain what it means to always wear face mask?”; “what it means to avoid touching the mouth/nose/eyes?”; “what does quarantine means?”; “what it means to avoid crowded places?”; “what it means to avoid travelling?”. This included questions for self-evaluation of compliance with the key measures considered in this study. The respondents were asked to grade themselves in a scale of 0% to 100%, where 0% meant that they did not comply at all with a specific measure, and 100% when they thought they complied completely with a given measure. Most of the questions had pre-defined answers for the interviewer to mark all the respondent’s answers (multiple options). The interviewers were instructed to ask the respondent “anything else?” repeatedly when the respondent stopped stating their responses, to explore the information as much as possible. There were open-ended questions and, for close-ended questions there were spaces for the interviewers to write all the responses that did not match the pre-defined options.

*<u>Implementation of COVID-19 preventive measures at household level</u> -*this included source of water to understand the presence and functioning of a handwashing facility, the capacity to keep 1.5 meters apart inside the household premises, number of face masks in the household; and compliance with quarantine for members or visitors who had contact with suspected cases. The fieldworkers were instructed to observe/verify the existence and functioning of a hand washing facility in the household (presence of water, soap and/or ashes), as recommend by the guidelines for collecting data on handwashing facilities (11). Ashes were recommended by the government for handwashing where there is no soap (13).

*<u>Symptoms of respiratory illness at household and community levels</u> –* the household heads were asked whether they had had any symptom of respiratory illness since 05 March 2020, when COVID-19 was first declared in South Africa; whether they or any member of the household had respiratory symptoms on the day of interview; and whether they knew about anyone with such symptoms in their neighbourhoods. The date when COVID-19 was first declared in South Africa was used as starting point because there were reports, on the media, of unprecedented inflow of Mozambicans returning from South Africa as the South African government had announced that would close the borders due to COVID-19; and this inflow was seen as a potential source for importing Coronavirus into Mozambique (5).

*<u>Perceived hardships due to COVID-19</u>* -the respondents were asked “in this problem of Coronavirus, what worries you the most?” The interviewers would listen to the respondent for mentions of any of the pre-defined options and pick all that were mentioned. The pre-defined options included: “Hunger”; “Travel restrictions”; “I stopped working”; “I am not working well”; “Afraid of being infected myself”; “Afraid that some family member may be infected”; “Coronavirus does not have cure”; “I am afraid to lose my job”; “I have lost my job”; “Interruptions of classes”; and others. Further, the survey asked whether there had been any social event that was postponed in the household due to Coronavirus

### 2.5. Concepts and methods of data analysis

The key COVID-19 preventive measures were defined according to the Manual for the Prevention of COVID-19 published by the Ministry of Health of Mozambique in April 2020 (13) and other standard definitions (14). Thus the wearing of face masks was defined as wearing a medical or non-medical mask by a person aged 6 years or more while indoor or outdoor settings where physical distancing of 1.5 meter minimum could not be maintained (15). Social distancing is keeping a minimum distance of 1.5 meter between people. Quarantine was defined as keeping someone who had a close contact with someone who has COVID-19 or is arriving from a place of risk of COVID-19, away from others for 14 days, including in their own home (16). Confinement or staying at home was defined as making all the efforts to stay at home and avoid travelling, leaving home only for cases of strong motives (13). Frequent hand washing was defined as washing hands with water and soap or disinfecting with alcohol every time that a person has contact with objects or other items that have or may have been touched by somebody else (13).

The analysis presents descriptive statistics on perceptions and levels of compliance with COVID-19 preventive measures at individual and at household levels, comparing the two districts and comparing by other characteristics such as rural/urban place of residence, occupation, age, and level of education. T-test was performed to assess the statistical significance of the differences on proportions observed between Manhiça and Quelimane or between categories of respondents in the same district. The open-ended responses and the texts written to specify the “other” category were analysed using content analysis (resorting to Stata *regexm* function, which performs a match of a word or expression in a text, to arrange them in groups according to the main content of each text), to see if they could fit in any of the pre-defined responses or new categories needed to be created. Vague responses that could not permit assessing the accuracy of respondent’s knowledge and could not fit in any of the created categories were classified as “incomprehensive or vague answer”. The analyses were done using Stata 14.2 (17).

## 3. RESULTS

### 3.1. Socio-demographic profile of participants and their households

The study interviewed 770 individuals, of which 62.3% were heads of households, 18.6%, their spouses, 11.0% their sons or daughters, and 8.1% others. Table 1 shows that 64.2% of participants were living in urban areas, with a higher percentage of urban residents in Quelimane (88.9%) than in Manhiça (39.3%), which is consistent with the urban-rural differences between the two study sites, as described earlier. The majority (65.7%) of the respondents were females, also with large differences between the two districts – more females in Manhiça (76.0%) than in Quelimane (55.4%), which is in line with the sex composition of the population in the Southern and Central regions of the country (more female-headed households in the South than in the Center), (18). By education, 17.5% of participants were illiterate, and 43.4% had only primary education, and 39.1% had secondary or higher education. In relation to occupation, the majority (68.4%) were unemployed and were engaged on subsistence family activities such as farming, fishing, production of fire wood/charcoal, followed by public sector officers and students (12.5%), and vendors in formal and informal sectors (9.5%). With regards to access to drinking water, 48.2% of households had their sources of water within the household premises, either piped water or wells, but Manhiça (a district that has only a small town) had higher percentage of households using piped water (59.6%) than Quelimane (36.8%), a district that has a major city) – probably because in Manhiça there are many private water suppliers who pump underground water through electric pumps and distribute by pipe to their clients, while in Quelimane the fewer private suppliers that exist do it by buying water from government sources and sell it in trucks, from one neighbourhood to another (19). Private suppliers may find it difficult to invest in water in Quelimane because the underground water is salty (19)

**Table 1.** Socio-demographic characteristics of participants by study area.

| Variable | Manhiça |  | Quelimane |  | Total |  |
| --- | --- | --- | --- | --- | --- | --- |
|  | n | % | n | % | N | % |
| **Total of participants** | 384 | 49.9 | 386 | 50.1 | 770 | 100 |
| **Relation to household head** |  |  |  |  |  |  |
| Head or spouse of head | 307 | 79.9 | 316 | 81.9 | 623 | 80.9 |
| Son or daughter | 35 | 9.1 | 50 | 12.9 | 85 | 11.0 |
| Other | 42 | 10.9 | 20 | 5.2 | 62 | 8.1 |
| **Sex of participant** |  |  |  |  |  |  |
| Male | 92 | 24.0 | 172 | 44.6 | 264 | 34.3 |
| Female | 292 | 76.5 | 214 | 55.4 | 506 | 65.7 |
| **Area of residence** |  |  |  |  |  |  |
| Urban | 151 | 39.3 | 343 | 88.9 | 494 | 64.2 |
| Rural | 233 | 60.7 | 43 | 11.1 | 276 | 35.8 |
| **Age group in years** |  |  |  |  |  |  |
| Adolescent (<19) | 15 | 3.9 | 22 | 5.7 | 37 | 4.8 |
| Adult (20 – 64) | 317 | 82.6 | 345 | 89.4 | 662 | 86.0 |
| Elder (65+) | 52 | 13.5 | 19 | 4.9 | 71 | 9.2 |
| **Level of education** |  |  |  |  |  |  |
| None | 93 | 24.2 | 42 | 10.9 | 135 | 17.5 |
| Primary | 196 | 51.0 | 138 | 35.8 | 334 | 43.4 |
| Secondary or higher | 95 | 24.7 | 206 | 53.4 | 301 | 39.1 |
| **Religion** |  |  |  |  |  |  |
| Protestant/Evangelic | 203 | 52.9 | 43 | 11.1 | 246 | 32.0 |
| Catholic | 31 | 8.1 | 218 | 56.5 | 249 | 32.6 |
| Zion/other/none | 144 | 37.5 | 30 | 7.8 | 174 | 22.6 |
| Islamic/Hindu | 6 | 1.6 | 95 | 24.6 | 101 | 13.6 |
| **Main occupation** |  |  |  |  |  |  |
| Peasants/fisher/charcoal/dn'twork | 285 | 74.5 | 242 | 62.7 | 527 | 68.4 |
| Teachers/public officers/studen | 24 | 6.3 | 72 | 18.7 | 96 | 12.5 |
| Vendors formal/informal | 25 | 6.5 | 48 | 12.4 | 73 | 9.5 |
| Carpenters/welding/builders | 50 | 13.0 | 24 | 6.2 | 74 | 9.6 |
| **Is your main source of water at home/yard** |  |  |  |  |  |  |
| Yes | 229 | 59.6 | 142 | 36.8 | 371 | 48.2 |
| No | 136 | 35.4 | 238 | 61.7 | 374 | 48.6 |
| Missing | 19 | 5.0 | 6 | 1.6 | 25 | 3.3 |
| **Main source of water, independent of location** |  |  |  |  |  |  |
| Piped water | 263 | 68.4 | 202 | 52.6 | 465 | 60.4 |
| Protected well | 19 | 5.0 | 48 | 12.5 | 67 | 8.7 |
| Unprotected well | 30 | 7.9 | 84 | 21.8 | 114 | 14.9 |
| Protected hole with pump | 56 | 14.6 | 3 | 0.8 | 59 | 7.7 |
| River, lake, rain, or other | 9 | 2.3 | 37 | 9.7 | 46 | 6.0 |
| Missing | 7 | 1.8 | 11 | 2.8 | 18 | 2.3 |

Source: Household survey, Manhiça 2020 and Quelimane, 2021.

### 3.2. Awareness about COVID-19 disease and its transmission dynamics

Table 2 presents the data on the awareness of COVID-19 disease by key socio-demographic characteristics of participants, and it shows that 98.7% of participants had heard of Coronavirus, but this percentage dropped to 89.2% when asked about COVID-19, which suggests that this disease was better known as Coronavirus than COVID-19 in October 2020 in Manhiça, and in February 2021 in Quelimane. The name COVID-19 was less known in Quelimane (86.0%) than in Manhiça (92.5%) (T-test *p-value = 0*.*014* when comparing the two proportions), and in urban areas (97.9%) than in rural areas (100%), although this rural-urban difference does not appear to be significant (*p=0*.*059*). Responses by religion appears to not differ between the two districts (*p=0*.*659*), but they do by occupation – public sector officers and students appear to be better informed than vendors and peasants/fisherman unemployed/retired (*p=0*.*005*). This is consistent with the expected, as one would expect public sector officers to be better informed about Coronavirus than peasants and fishermen, as they tend to have more access to the media and other means of public communication. In both Manhiça and Quelimane, the awareness about Coronavirus disease decreased with age and increased with level of education, and although this variation was not statistically significant in both Manhica (*p=0*.*826*) and Quelimane (*p=0*.*963*) it appears to be an expected pattern. More than a third (34.3%) of those who said they had heard of Coronavirus or COVID-19 said they were unable to define Coronavirus, 18.7% said that it was a disease caused by a virus, 14.8% said it was a respiratory disease, 10.0%, a disease that causes cough; 9.0%, dangerous, deadly incurable disease; and 4.6% a disease that comes from China.

**Table 2.** Awareness about COVID-19 disease by socio-demographic characteristics of participants, Manhiça and Quelimane.

| Variable | Manhiça |  | Quelimane |  | Total |  | Chi2<br>P-value<sup>1</sup> |
| --- | --- | --- | --- | --- | --- | --- | --- |
|  | n | % | n | % | N | % |  |
| Total of participants | 384 | 49.9 | 386 | 50.1 | 770 | 100 |  |
| PANEL “A” |  |  |  |  |  |  |  |
| Have you ever heard of Coronavirus |  |  |  |  |  |  |  |
| Yes | 379 | 98.7 | 381 | 98.7 | 760 | 98.7 | 0.788 |
| No | 3 | 0.8 | 4 | 1.0 | 7 | 0.9 |  |
| Missing | 2 | 0.5 | 1 | 0.3 | 3 | 0.4 |  |
| Have you ever heard of COVID-19 |  |  |  |  |  |  |  |
| Yes | 355 | 92.5 | 332 | 86.0 | 687 | 89.2 | 0.014 |
| No | 28 | 7.3 | 53 | 13.7 | 81 | 10.5 |  |
| Missing | 1 | 0.3 | 1 | 0.3 | 2 | 0.3 |  |
| PANEL “B” |  |  |  |  |  |  |  |
| Awareness of Coronavirus by selected characteristics: % of those who said YES<sup>2</sup> |  |  |  |  |  |  |  |
| Place of residence |  |  |  |  |  |  |  |
| Urban | 146 | 96.7<sup>3</sup> | 338 | 98.5 | 494 | 97.9 | 0.059 |
| Rural | 233 | 100.0 | 43 | 100.0 | 276 | 100.0 |  |
| Age group in years |  |  |  |  |  |  |  |
| Adolescent (<20) | 15 | 100.0 | 22 | 100.0 | 37 | 100.0 | 0.906 |
| Adult (20 – 64) | 313 | 98.7 | 340 | 98.6 | 653 | 98.6 |  |
| Elder (65+) | 51 | 98.1 | 19 | 100.0 | 70 | 98.6 |  |
| Level of education |  |  |  |  |  |  |  |
| None | 91 | 97.9 | 41 | 97.6 | 132 | 97.8 | 0.106 |
| Primary | 193 | 98.5 | 135 | 97.8 | 328 | 98.2 |  |
| Secondary and more | 95 | 100.0 | 205 | 99.5 | 300 | 99.6 |  |
| Religion |  |  |  |  |  |  |  |
| Catholic | 31 | 100.0 | 215 | 98.6 | 246 | 98.8 | 0.659 |
| Protestant/Evangelic | 201 | 99.0 | 43 | 100.0 | 244 | 99.2 |  |
| Zion/none/others/none | 141 | 97.9 | 29 | 96.7 | 170 | 97.7 |  |
| Islamic/Hindu | 6 | 100.0 | 94 | 99.0 | 100 | 99.0 |  |
| Main occupation |  |  |  |  |  |  |  |
| Peasants/fisher/unemployed | 282 | 99.0 | 241 | 99.6 | 523 | 99.2 | 0.005 |
| Vendors formal/informal | 24 | 96.0 | 45 | 93.8 | 69 | 94.5 |  |
| Carpenters/welding/builders | 49 | 98.0 | 23 | 95.8 | 72 | 97.3 |  |
| Public sector officers/ students | 24 | 100.0 | 72 | 100.0 | 96 | 100.0 |  |

Source: Household survey, Manhiça 2020 and Quelimane, 2021.
<sup>1</sup> The P-value in Panel "A" and "B" refers to Pearson X<sup>2</sup> test of how significant are the differences of responses between Manhiça and Quelimane.
<sup>2</sup> The focus on those who said YES do Coronavirus is because only 3 people said YES to both Coronavirus and Covid-19, so there is no need to repeat the tabulations for only 3 respondents.
<sup>3</sup> Chi<sup>2</sup> P-value in Manhiça is = 0.009.

Table 3 presents the results on the knowledge of symptoms, means of transmission and prevention of Coronavirus, and it shows that dry cough (17.8%), fever (15.7%), flu-like symptoms (14.2%), breathing difficulties (13.6%), and pain in the throat (8.8%) were the symptoms most mentioned by the respondents. By district, significant differences were observed in relation to the proportion of respondents who mentioned fever (*p=0*.*003*), flu-like symptoms (*p=0*.*000*), pain in the throat (*p=0*.*002*), and those who said did not know (*p=0*.*000*). The mechanisms of transmission most mentioned are touching infected person or object (30.9%), inhaling the air from the mouth of an infected person (21.3%), contact with saliva droplets from an infected person (20.4%), and touching the mouth, nose or eyes (13.3%). The means of prevention most mentioned were washing hands with soap or alcohol (30.9%), using face mask always (28.6%), social distancing (16.6%), and avoiding crowded places (10.2%). By district significant differences were observed in relation to avoiding crowded places (*p=0*.*000*) and avoiding travelling (*p=0*.*000*).

**Table 3.** : Knowledge of symptoms, means of transmission and prevention of Coronavirus

| Variable | Manhiça |  | Quelimane |  | Total |  | Chi2 |
| --- | --- | --- | --- | --- | --- | --- | --- |
|  | n | %<sup>4</sup> | n | %<sup>7</sup> | N | %<sup>5</sup> | P-value |
| **What are the symptoms of Coronavirus? (multiple options) [If have heard of Coronavirus]** |  |  |  |  |  |  |  |
| Dry cough | 184 | 47.9 | 201 | 52.1 | 385 | 17.8 | 0.249 |
| Fever | 149 | 38.8 | 191 | 49.5 | 340 | 15.7 | 0.003 |
| Flu-like symptoms | 183 | 47.7 | 124 | 32.1 | 307 | 14.2 | 0.000 |
| Breathing difficulties | 142 | 37.0 | 153 | 39.6 | 295 | 13.6 | 0.448 |
| Head ache | 124 | 32.3 | 159 | 41.2 | 283 | 13.1 | 0.010 |
| Pain in the throat | 113 | 29.4 | 76 | 19.7 | 189 | 8.7 | 0.002 |
| Cough with sputum | 58 | 15.1 | 68 | 17.6 | 126 | 5.8 | 0.346 |
| Muscular pain | 61 | 15.9 | 60 | 15.7 | 121 | 5.6 | 0.896 |
| Don't know | 12 | 3.1 | 43 | 11.1 | 55 | 2.5 | 0.000 |
| Vomiting | 20 | 5.2 | 30 | 7.8 | 50 | 2.3 | 0.149 |
| Other | 7 | 1.8 | 7 | 1.8 | 14 | 0.6 | 0.992 |
| Total multiple responses | 1,053 | - | 1,112 | - | 2,165 | 100 | - |
| **How is Coronavirus transmitted? (multiple options)** |  |  |  |  |  |  |  |
| Touch infected person/object<sup>6</sup> | 240 | 62.5 | 248 | 64.3 | 488 | 30.9 | 0.615 |
| Inhale air mouth of infected | 186 | 48.4 | 150 | 38.9 | 336 | 21.3 | 0.007 |
| Droplets of saliva of infected | 169 | 44.0 | 153 | 39.6 | 322 | 20.4 | 0.219 |
| Touch mouth or nose, eyes | 106 | 27.6 | 104 | 26.9 | 210 | 13.3 | 0.837 |
| Kiss infected person | 100 | 26.0 | 61 | 15.8 | 161 | 10.2 | 0.000 |
| Other transmission means | 16 | 4.2 | 44 | 11.4 | 60 | 3.8 | 0.000 |
| Do not know | 0 | 0.0 | 2 | 0.5 | 2 | 0.1 | 0.158 |
| Total of multiple responses | 817 | - | 762 | - | 1,500 | 100.0 | - |
| **How can we prevent Coronavirus? (multiple options)** |  |  |  |  |  |  |  |
| Wash hand/soap or alcohol | 298 | 77.6 | 318 | 82.4 | 616 | 30.9 | 0.097 |
| Use face mask always | 275 | 71.6 | 296 | 76.7 | 571 | 28.6 | 0.108 |
| Social distancing | 155 | 40.4 | 177 | 45.9 | 332 | 16.6 | 0.124 |
| Avoid crowded places | 124 | 32.4 | 80 | 20.7 | 204 | 10.2 | 0.000 |
| Avoid touch mouth/noise/eye | 57 | 14.8 | 53 | 13.7 | 110 | 5.5 | 0.659 |
| Other means | 21 | 5.5 | 42 | 10.9 | 63 | 3.2 | 0.006 |
| Quarantine | 31 | 8.1 | 23 | 6.0 | 54 | 2.7 | 0.251 |
| Avoid travelling | 42 | 10.9 | 3 | 0.8 | 45 | 2.3 | 0.000 |
| Total multiple responses | 1,003 | - | 992 | - | 1,995 | 100.0 | - |

Source: Household survey, Manhiça 2020 and Quelimane, 2021.
<sup>4</sup> % of respondents that mentioned a symptom – tells how many respondents mentioned a symptom
<sup>5</sup> % of times that a symptom was mentioned – tells which symptom was most mentioned.
<sup>6</sup> Including hugging infected person

### 3.3. Knowledge and perceptions of COVID-19 preventive measures

Table 4 presents the results on how people understand the COVID-19 preventive measures. The majority of respondents knew that social distancing refers to keeping some distance between each other (62.0%) [62.0% comes from summing “keeping 1.5-2 meters from others” (43.9%) and “to be away/be distant from others” (18.1%)], but 19.9% gave incomprehensive or vague answers, and 17.5% said that they did not know. In relation to hand washing in the context of COVID-19, 32.9% of respondents gave vague responses, 28.6% said that they wash their hands when leaving or arriving at home; 26.0%, after touching something or someone; and 7.7%, after using the toilet or when the hands are dirty or when they are about to eat something. Quarantine was defined as staying at home or in the same place by 36.6% of respondents; 26.5% defined it comprehensively as isolating someone who is sick or suspected or who had contact with positive or suspected cases (some respondents in this group indicated the period of 14 days); but 29.2% said that they did not know what is was.

**Table 4.** Knowledge and perceptions of the recommend anti-COVID-19 measures

| Variable | Manhiça |  | Quelimane |  | Total |  | Chi2 |
| --- | --- | --- | --- | --- | --- | --- | --- |
|  | n | % | n | % | N | % | P-value |
| **Total of participants** | 384 | 49.9 | 386 | 50.1 | 770 | 100 |  |
| **What social distancing means?** |  |  |  |  |  |  |  |
| To keep 1,5-2m from others | 170 | 44.3 | 168 | 43.5 | 338 | 43.9 | 0.000 |
| To be away/distant from other | 37 | 9.6 | 102 | 26.4 | 139 | 18.1 |  |
| Inconclusive/astray answer | 93 | 24.2 | 60 | 15.5 | 153 | 19.9 |  |
| Don't know | 79 | 20.6 | 56 | 14.5 | 135 | 17.5 |  |
| Missing | 5 | 1.3 | 0 | 0.0 | 5 | 0.6 |  |
| **In which situations do you wash your hands, in this context of COVID-19** |  |  |  |  |  |  |  |
| Inconclusive/astray answer | 173 | 45.1 | 80 | 20.7 | 253 | 32.9 | 0.000 |
| When leaving/returning home | 47 | 12.2 | 173 | 44.8 | 220 | 28.6 |  |
| After touching something/one | 116 | 30.2 | 84 | 21.8 | 200 | 26.0 |  |
| After toilet/before meals/dirty | 14 | 3.6 | 45 | 11.7 | 59 | 7.7 |  |
| Don't know | 24 | 6.3 | 3 | 0.8 | 27 | 3.5 |  |
| Missing | 10 | 2.6 | 1 | 0.3 | 11 | 1.4 |  |
| **What it means to avoid crowded places?** |  |  |  |  |  |  |  |
| Inconclusive/astray answer | 113 | 29.4 | 95 | 24.6 | 208 | 27.0 | 0.000 |
| Not more than 5-10 people | 90 | 23.4 | 103 | 26.7 | 193 | 25.1 |  |
| Many people | 30 | 7.8 | 109 | 28.2 | 139 | 18.1 |  |
| Not more than 20 people | 57 | 14.8 | 23 | 6.0 | 80 | 10.4 |  |
| Don't know | 31 | 8.1 | 42 | 10.9 | 73 | 9.5 |  |
| Not more than 50 people | 43 | 11.2 | 7 | 1.8 | 50 | 6.5 |  |
| Missing | 20 | 5.2 | 7 | 1.8 | 27 | 3.5 |  |
| **What it means to avoid travelling?** |  |  |  |  |  |  |  |
| Inconclusive/astray answer | 208 | 54.2 | 291 | 75.4 | 499 | 64.8 | 0.000 |
| Don't know | 67 | 17.4 | 59 | 15.3 | 126 | 16.4 |  |
| To stay at home | 58 | 15.1 | 25 | 6.5 | 83 | 10.8 |  |
| To travel only for extreme case | 42 | 10.9 | 2 | 0.5 | 44 | 5.7 |  |
| Missing | 9 | 2.3 | 9 | 2.3 | 18 | 2.3 |  |
| **What it means quarantine?** |  |  |  |  |  |  |  |
| To stay at home or same place | 121 | 31.5 | 161 | 41.7 | 282 | 36.6 | 0.000 |
| Don't know | 112 | 29.2 | 113 | 29.3 | 225 | 29.2 |  |
| To isolate sick/suspect person | 127 | 33.1 | 77 | 19.9 | 204 | 26.5 |  |
| Inconclusive/astray answer | 14 | 3.6 | 25 | 6.5 | 39 | 5.1 |  |
| Missing | 4 | 1.0 | 10 | 2.6 | 14 | 1.8 |  |
| **What it means to avoid touching the mouth, or eyes, or nose** |  |  |  |  |  |  |  |
| Inconclusive/astray answer | 112 | 29.2 | 210 | 54.4 | 322 | 41.8 | 0.000 |
| Wash hands before touch them | 147 | 38.3 | 60 | 15.5 | 207 | 26.9 |  |
| Don't know | 72 | 18.8 | 83 | 21.5 | 155 | 20.1 |  |
| Missing | 53 | 13.8 | 33 | 8.5 | 86 | 11.2 |  |

378 Source: Household survey, Manhiça 2020 and Quelimane, 2021.

### 3.4. Implementation of anti-COVID-19 measures at individual level

Table 5 presents the results of participants’ self-assessment of their level of compliance with anti-COVID-19 measures. It shows that 35.8% of the respondents said that they could not stay at home at all, and only 20.0% could stay at home 100% of the time. Only 21.9% of respondents said that they could wash their hands on all occasions thought to be necessary. Mask wearing in public places appears to be the measure that more people (48.8%) think they complied with it in 100% of the occasions, followed by avoiding unnecessary travels (40.0%), avoiding crowed places (34.0%), and social distancing outside the household (34.4%). On the other hand, the most difficult anti-COVID-19 measure appears to be avoiding touching the mouth or nose or eyes (with 77.4% of respondents saying that they did not comply with it or complied only in 25% or 50% of the occasions (77.4% is the sum of 28.7%, 25.1% and 23.6%). This is followed by social distancing within the household (73.1%), staying at home (66.2%) and frequent hand washing (56.0%).

**Table 5.** Self-assessment of compliance with anti-COVID-19 measures at individual level

| Variable | Manhiça |  | Quelimane |  | Total |  | Chi2<br>P-value |
| --- | --- | --- | --- | --- | --- | --- | --- |
|  | n | % | n | % | N | % |  |
| **Total of participants** | 384 | 49.9 | 386 | 50.1 | 770 | 100 |  |
| **Degree of compliance with staying at home** |  |  |  |  |  |  |  |
| 0% | 153 | 39.8 | 123 | 31.9 | 276 | 35.8 | 0.000 |
| 100% | 98 | 25.5 | 56 | 14.5 | 154 | 20.0 |  |
| 25% | 57 | 14.8 | 63 | 16.3 | 120 | 15.6 |  |
| 50% | 44 | 11.5 | 70 | 18.1 | 114 | 14.8 |  |
| 75% | 23 | 6.0 | 74 | 19.2 | 97 | 12.6 |  |
| **Degree of compliance with hand-washing always** |  |  |  |  |  |  |  |
| 50% | 132 | 34.4 | 94 | 24.4 | 226 | 29.4 | 0.000 |
| 25% | 53 | 13.8 | 119 | 30.8 | 172 | 22.3 |  |
| 100% | 107 | 27.9 | 62 | 16.1 | 169 | 21.9 |  |
| 75% | 87 | 22.7 | 80 | 20.7 | 167 | 21.7 |  |
| 0% | 2 | 0.5 | 31 | 8.0 | 33 | 4.3 |  |
| **Degree of compliance with face mask wearing in public places** |  |  |  |  |  |  |  |
| 100% | 192 | 50.0 | 184 | 47.7 | 376 | 48.8 | 0.000 |
| 50% | 85 | 22.1 | 87 | 22.5 | 172 | 22.3 |  |
| 75% | 70 | 18.2 | 71 | 18.4 | 141 | 18.3 |  |
| 25% | 32 | 8.3 | 39 | 10.1 | 71 | 9.2 |  |
| 0% | 3 | 0.8 | 5 | 1.3 | 8 | 1.0 |  |
| **Degree of compliance with avoiding touching the mouth, nose, and eyes** |  |  |  |  |  |  |  |
| 0% | 18 | 4.7 | 203 | 52.6 | 221 | 28.7 |  |
| 25% | 131 | 34.1 | 62 | 16.1 | 193 | 25.1 |  |
| 50% | 129 | 33.6 | 53 | 13.7 | 182 | 23.6 | 0.000 |
| 75% | 52 | 13.5 | 60 | 15.5 | 112 | 14.5 |  |
| 100% | 52 | 13.5 | 7 | 1.8 | 59 | 7.7 |  |
| **Degree of compliance with social distancing within the household** |  |  |  |  |  |  |  |
| 0% | 18 | 4.7 | 192 | 49.7 | 210 | 27.3 |  |
| 25% | 131 | 34.1 | 51 | 13.2 | 182 | 23.6 |  |
| 50% | 129 | 33.6 | 42 | 10.9 | 171 | 22.2 | 0.000 |
| 75% | 52 | 13.5 | 50 | 13.0 | 102 | 13.2 |  |
| 100% | 52 | 13.5 | 7 | 1.8 | 59 | 7.7 |  |
| **Degree of compliance with social distancing outside the household** |  |  |  |  |  |  |  |
| 75% | 111 | 28.9 | 154 | 39.9 | 265 | 34.4 |  |
| 100% | 138 | 35.9 | 85 | 22.0 | 223 | 29.0 |  |
| 50% | 94 | 24.5 | 93 | 24.1 | 187 | 24.3 | 0.000 |
| 25% | 31 | 8.1 | 37 | 9.6 | 68 | 8.8 |  |
| 0% | 8 | 2.1 | 17 | 4.4 | 25 | 3.2 |  |
| **Degree of compliance with avoiding crowded places** |  |  |  |  |  |  |  |
| 100% | 157 | 40.9 | 105 | 27.2 | 262 | 34.0 |  |
| 75% | 96 | 25.0 | 128 | 33.2 | 224 | 29.1 |  |
| 50% | 94 | 24.5 | 99 | 25.6 | 193 | 25.1 | 0.000 |
| 25% | 30 | 7.8 | 33 | 8.5 | 63 | 8.2 |  |
| 0% | 5 | 1.3 | 21 | 5.4 | 26 | 3.4 |  |
| **Degree of compliance with avoiding unnecessary travels** |  |  |  |  |  |  |  |
| 100% | 208 | 54.2 | 100 | 25.9 | 308 | 40.0 |  |
| 50% | 68 | 17.7 | 99 | 25.6 | 167 | 21.7 |  |
| 75% | 79 | 20.6 | 62 | 16.1 | 141 | 18.3 | 0.000 |
| 25% | 19 | 4.9 | 95 | 24.6 | 114 | 14.8 |  |
| 0% | 8 | 2.1 | 30 | 7.8 | 38 | 4.9 |  |

Source: Household survey, Manhiça 2020 and Quelimane, 2021.

### 3.5. Implementation of anti-COVID-19 measures at household level

Table 6 presents the degree of compliance with anti-COVID-19 measures at household level, combining data from questions and observations as described in the section on methods. It shows that sixty-nine per cent (69.4%) of households did not have hand washing facilities or disinfection (75.6% in Quelimane and 63.0% in Manhiça); and of those that did have devices, 58.5% had no water in the hand washing facility, 62.0% had no soap, and 74.4% had no ash in these devices. In addition to hand washing facilities, it was asked whether there was any other disinfectant or not – 22.6% had at least one other disinfectant such as alcohol or alcohol gel. The other indicator was the existence and the number of masks in the household -almost all households (98.1%) had masks, but the ratio of masks per household member was very low (median of 1 mask per member aged 6+ years), which suggests that people were using the same mask for too long time without replacing it every few hours as recommended by the Ministry of Health of Mozambique (13).

**Table 6.**
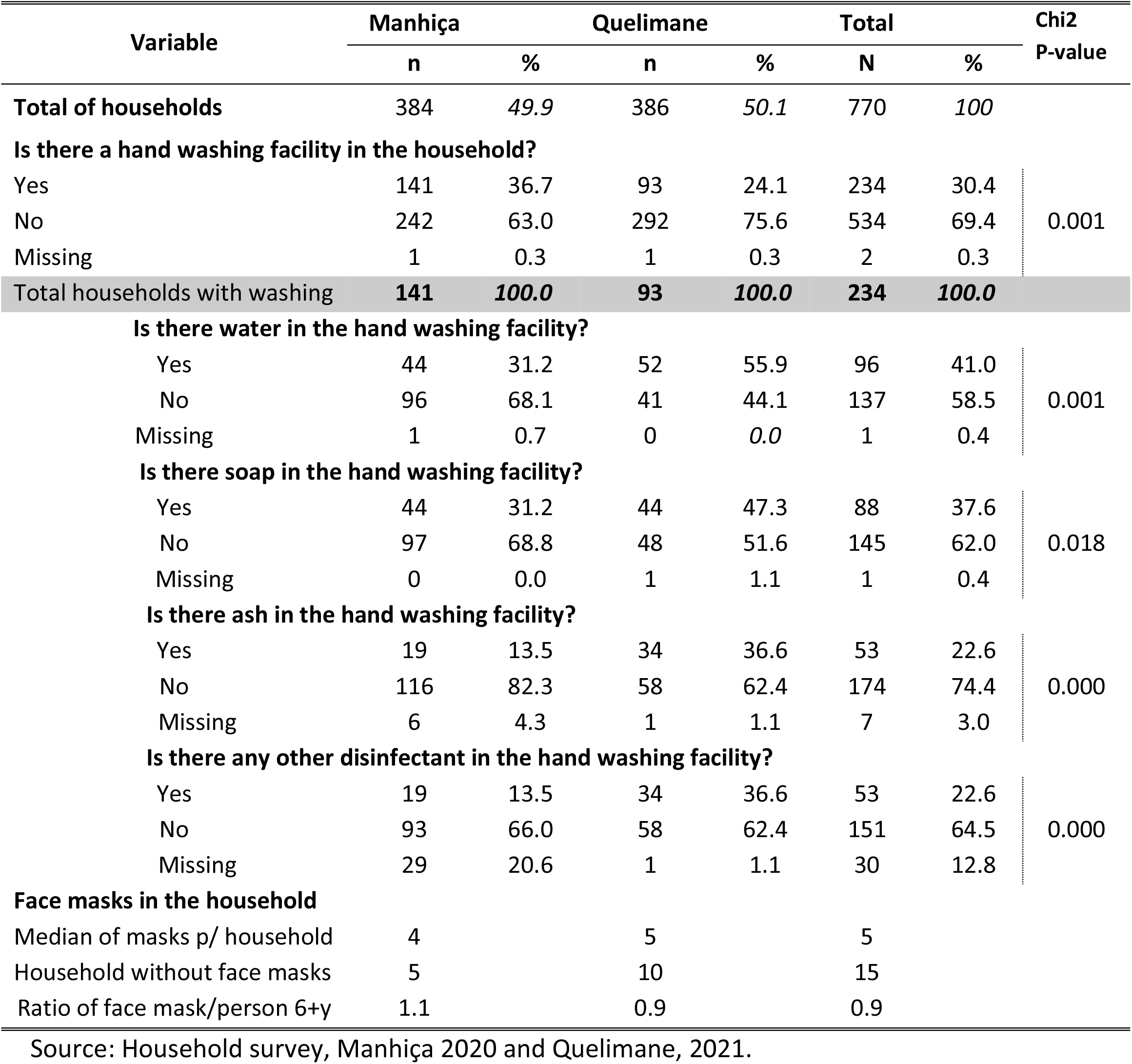
Implementation of anti-COVID-19 measures at household level, Manhiça and Quelimane

| Variable | Manhiça |  | Quelimane |  | Total |  | Chi2<br>P-value |
| --- | --- | --- | --- | --- | --- | --- | --- |
|  | n | % | n | % | N | % |  |
| **Total of households** | 384 | 49.9 | 386 | 50.1 | 770 | 100 |  |
| **Is there a hand washing facility in the household?** |  |  |  |  |  |  |  |
| Yes | 141 | 36.7 | 93 | 24.1 | 234 | 30.4 | 0.001 |
| No | 242 | 63.0 | 292 | 75.6 | 534 | 69.4 |  |
| Missing | 1 | 0.3 | 1 | 0.3 | 2 | 0.3 |  |
| **Total households with washing** | **141** | **100.0** | **93** | **100.0** | **234** | **100.0** |  |
| **Is there water in the hand washing facility?** |  |  |  |  |  |  |  |
| Yes | 44 | 31.2 | 52 | 55.9 | 96 | 41.0 | 0.001 |
| No | 96 | 68.1 | 41 | 44.1 | 137 | 58.5 |  |
| Missing | 1 | 0.7 | 0 | 0.0 | 1 | 0.4 |  |
| **Is there soap in the hand washing facility?** |  |  |  |  |  |  |  |
| Yes | 44 | 31.2 | 44 | 47.3 | 88 | 37.6 | 0.018 |
| No | 97 | 68.8 | 48 | 51.6 | 145 | 62.0 |  |
| Missing | 0 | 0.0 | 1 | 1.1 | 1 | 0.4 |  |
| **Is there ash in the hand washing facility?** |  |  |  |  |  |  |  |
| Yes | 19 | 13.5 | 34 | 36.6 | 53 | 22.6 | 0.000 |
| No | 116 | 82.3 | 58 | 62.4 | 174 | 74.4 |  |
| Missing | 6 | 4.3 | 1 | 1.1 | 7 | 3.0 |  |
| **Is there any other disinfectant in the hand washing facility?** |  |  |  |  |  |  |  |
| Yes | 19 | 13.5 | 34 | 36.6 | 53 | 22.6 | 0.000 |
| No | 93 | 66.0 | 58 | 62.4 | 151 | 64.5 |  |
| Missing | 29 | 20.6 | 1 | 1.1 | 30 | 12.8 |  |
| **Face masks in the household** |  |  |  |  |  |  |  |
| Median of masks p/ household | 4 |  | 5 |  | 5 |  |  |
| Household without face masks | 5 |  | 10 |  | 15 |  |  |
| Ratio of face mask/person 6+y | 1.1 |  | 0.9 |  | 0.9 |  |  |

Source: Household survey, Manhiça 2020 and Quelimane, 2021.

### 3.6. Prevalence of symptoms of respiratory illness

The results on symptoms of respiratory illness show that 93 (12.1%) respondents have had symptoms sometime since COVID-19 was announced first in South Africa (05 March 2020), and 38.1% of them had symptoms in two or more episodes. However, only 6 households (0.8%) had a person with symptoms at the date of interview, of which 4 had only one person and 2 had two people with symptoms. By district, Quelimane had significantly higher number of (past and current) cases with symptoms 80 (20.7% of respondents) than Manhiça 13 (3.4%), (*p=0*.*000*).

### 3.7. Perceived hardships due to Coronavirus

The results on the hardships felt by the population due to Coronavirus show that the closure of educational institutions was the leading hardship (with 18.3% of mentions), followed by hunger (17.5%), fear of being infected or having someone from the family infected (12.2%), and other worries (12.0%), (Table 7). The “other worries” included fear of death and of uncertainty of the future (32.1%), closure of churches (28.3%), and loss of opportunities for employment and other means of livelihood (18.3%). Most of these hardships were reported with significant differences by districts (see those with p-value < 0.05).

**Table 7.**
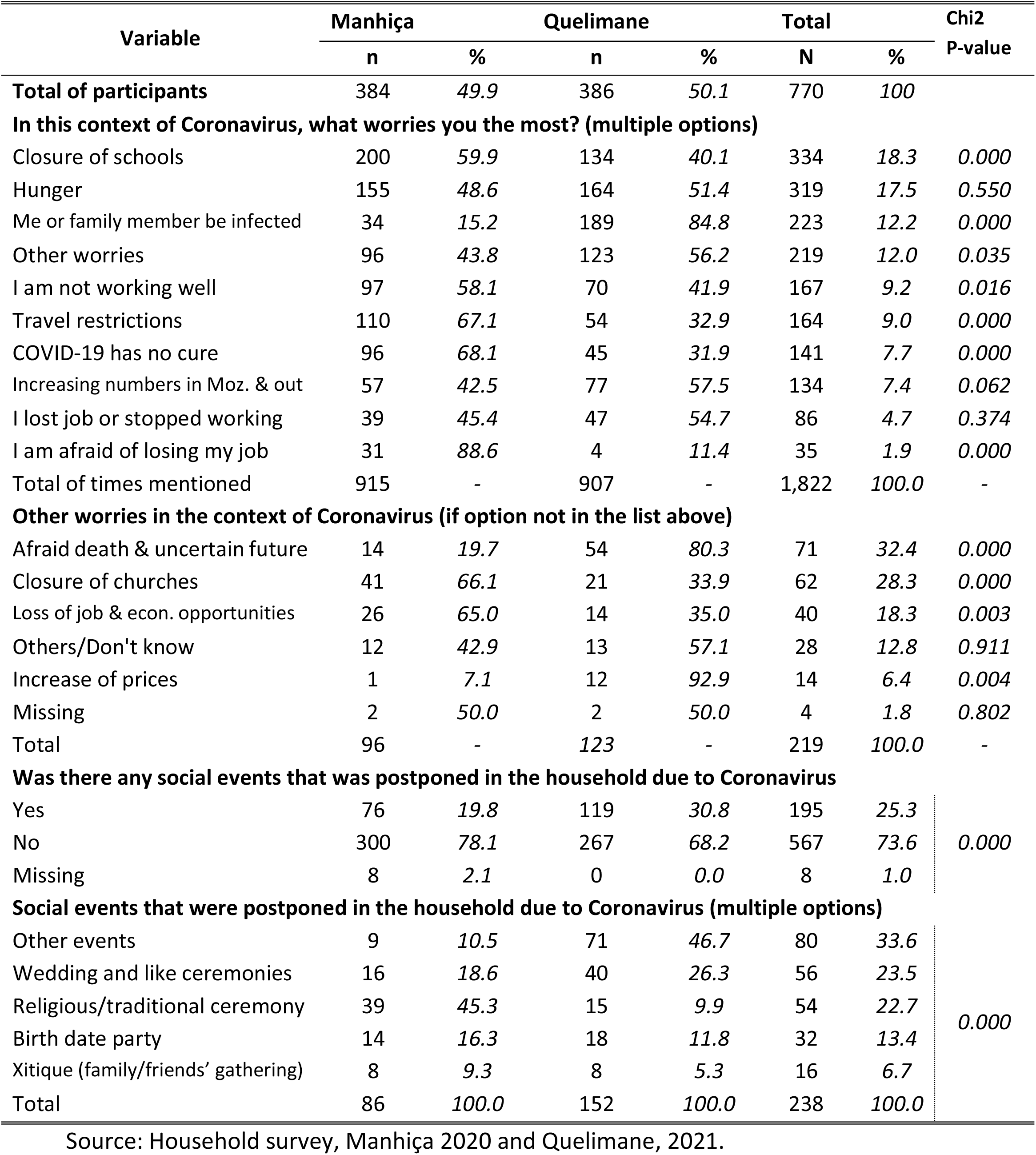
Participants’ worries caused by Coronavirus in Manhiça and Quelimane

| Variable | Manhiça |  | Quelimane |  | Total |  | Chi2 |
| --- | --- | --- | --- | --- | --- | --- | --- |
|  | n | % | n | % | N | % | P-value |
| Total of participants | 384 | 49.9 | 386 | 50.1 | 770 | 100 |  |
| In this context of Coronavirus, what worries you the most? (multiple options) |  |  |  |  |  |  |  |
| Closure of schools | 200 | 59.9 | 134 | 40.1 | 334 | 18.3 | 0.000 |
| Hunger | 155 | 48.6 | 164 | 51.4 | 319 | 17.5 | 0.550 |
| Me or family member be infected | 34 | 15.2 | 189 | 84.8 | 223 | 12.2 | 0.000 |
| Other worries | 96 | 43.8 | 123 | 56.2 | 219 | 12.0 | 0.035 |
| I am not working well | 97 | 58.1 | 70 | 41.9 | 167 | 9.2 | 0.016 |
| Travel restrictions | 110 | 67.1 | 54 | 32.9 | 164 | 9.0 | 0.000 |
| COVID-19 has no cure | 96 | 68.1 | 45 | 31.9 | 141 | 7.7 | 0.000 |
| Increasing numbers in Moz. & out | 57 | 42.5 | 77 | 57.5 | 134 | 7.4 | 0.062 |
| I lost job or stopped working | 39 | 45.4 | 47 | 54.7 | 86 | 4.7 | 0.374 |
| I am afraid of losing my job | 31 | 88.6 | 4 | 11.4 | 35 | 1.9 | 0.000 |
| Total of times mentioned | 915 | - | 907 | - | 1,822 | 100.0 | - |
| Other worries in the context of Coronavirus (if option not in the list above) |  |  |  |  |  |  |  |
| Afraid death & uncertain future | 14 | 19.7 | 54 | 80.3 | 71 | 32.4 | 0.000 |
| Closure of churches | 41 | 66.1 | 21 | 33.9 | 62 | 28.3 | 0.000 |
| Loss of job & econ. opportunities | 26 | 65.0 | 14 | 35.0 | 40 | 18.3 | 0.003 |
| Others/Don't know | 12 | 42.9 | 13 | 57.1 | 28 | 12.8 | 0.911 |
| Increase of prices | 1 | 7.1 | 12 | 92.9 | 14 | 6.4 | 0.004 |
| Missing | 2 | 50.0 | 2 | 50.0 | 4 | 1.8 | 0.802 |
| Total | 96 | - | 123 | - | 219 | 100.0 | - |
| Was there any social events that was postponed in the household due to Coronavirus |  |  |  |  |  |  |  |
| Yes | 76 | 19.8 | 119 | 30.8 | 195 | 25.3 | 0.000 |
| No | 300 | 78.1 | 267 | 68.2 | 567 | 73.6 |  |
| Missing | 8 | 2.1 | 0 | 0.0 | 8 | 1.0 |  |
| Social events that were postponed in the household due to Coronavirus (multiple options) |  |  |  |  |  |  |  |
| Other events | 9 | 10.5 | 71 | 46.7 | 80 | 33.6 | 0.000 |
| Wedding and like ceremonies | 16 | 18.6 | 40 | 26.3 | 56 | 23.5 |  |
| Religious/traditional ceremony | 39 | 45.3 | 15 | 9.9 | 54 | 22.7 |  |
| Birth date party | 14 | 16.3 | 18 | 11.8 | 32 | 13.4 |  |
| Xitique (family/friends' gathering) | 8 | 9.3 | 8 | 5.3 | 16 | 6.7 |  |
| Total | 86 | 100.0 | 152 | 100.0 | 238 | 100.0 |  |

Source: Household survey, Manhiça 2020 and Quelimane, 2021.

In relation to postponement of social events due to Coronavirus, 25.3% of respondents said that they had postponed at least one social event in their household – 31.8% of these households postponed more than one event. Wedding and alike ceremonies (23.5%), religious and traditional ceremonies (22.7%), birth date parties (13.4%), and gatherings for *xitique* (6.7%) are the leading postponed events. *Xitique* is a term in local language of Southern Mozambique (Changana), but also used equally in Quelimane, for a network or group of friends, family members, or colleagues for inter-support or other means of solidarity involving money saving which is rotatively paid to one member of the group. Often it involves social gathering in the household of the member that is receiving the group’s savings (20), (21).

## 4. DISCUSSION

This study aimed to assess community perceptions and levels of compliance with anti-COVID-19 measures recommended by the government of Mozambique in the districts of Manhiça and Quelimane. At the time the study was designed and implemented, there were several challenges both from the conceptual and operational perspectives, as well as from the safety point of view, as both the researchers and the participants were afraid of exposure to a very contagious and deadly disease by conducting or participating in the study. From the perspectives of concepts and operational procedures, some concepts were new to the general public, and most importantly, some concepts were changing over the course of the pandemic, for example, the term “social distancing” has changed to “physical distancing”, and “crowed places” were venue- and time-variant and the maximum number of people allowed to gather together in one place changed continually as the disease was evolving. Also, the study required visiting hundreds of households (with more than 3,500 people) to study a disease that had forced people to refrain from moving or visiting each other to avoid transmission. Even the IRB at CISM was puzzled on whether this study could be approved due to safety issues and the request made by the study team for using oral audio-recorded consents instead of the commonly used written consents. At that time, most studies were collecting data using information and communication technologies (ICTs)(22), but this was not recommended for Manhiça and Quelimane where the coverage of ICTs was very limited – which could lead to exclusion of participants without access to ICTs (23). For example, a study on COVID-19 preventive behaviour using ICTs in China, required that participants should have been previously registered in an online-based survey platform and be literate, which means that all those not registered in that platform or who were illiterate were excluded; and it ended up with only 13% of participants from rural areas and 87% from urban areas (24).

Given the absence of previous studies on how community members understand and comply with anti-COVID-19 measures in Mozambique and other countries, designing the questionnaires was very challenging. Thus, the research team resorted to open-ended questions for the interviewers to write everything the respondent would say because there was no literature to guide the elaboration of pre-defined responses. However, too many open-ended questions (and too many “others, specify”) led to hard work during the analysis than would have been if the majority of questions were close-ended. This led to delay in the analysis and publication of the results, although it served also as a good opportunity to capture as much as possible variability of responses than would otherwise have been; and it improved the researchers’ ability to analysis lengthy text-responses in a quantitative survey.

The results show that most participants had heard of Coronavirus and COVID-19 by the time of the survey, with more people acknowledging Coronavirus than COVID-19, both in Manhiça and in Quelimane. At that time, there were too many new concepts around the pandemic and the terms Coronavirus and COVID-19 appeared to be used interchangeably to mean the same disease. Even the term “novel Coronavirus” was not as popular as the two, let alone the term SARS-CoV-2. Likewise, even some apparently obvious terms like confinement, stay at home, immobility, and quarantine had not been well communicated to the public in Mozambique. This leads to a recommendation that in public emergencies such as these, policy makers should be cautious to use the same terms consistently, and avoid similar but not equal terms, for example, it has never been very clear whether confinement, isolation, staying at home, and immobility meant the same or they were different within the context of Coronavirus. Later on, a new term started to be publically used without a clear definition, e.g. “the new normal” – intended to advise the population to adapt to this new context for longer periods of time -but none knows how this was interpreted because sometimes it was used simultaneously with relaxation of anti-COVID-19 measures. Further, at the time this survey was conducted, some respondents in rural and urbans areas of Central Mozambique did not believe that COVID-19 existed, and others said that fighting for food was of high priority than following anti-COVID-19 measures (Chaimite, 2020); and in Maputo city, misinformation was considered an important issue for policy makers to deal with (PERC, 2020) -this study showed that 72% of respondents believed hot climate prevents COVID-19, 41% believed COVID-19 is a germ weapon created by a government; and 39% said they would like more information, particularly on COVID-19 protection, causes and cure (PERC, 2020).

This study shows highs percentages of vague or inconclusive responses (ranging from 5.1% in the definition of quarantine, to 64.8% in the definition of avoiding travelling). Adding the “don’t knows” to these vague responses leads to the conclusion that most respondents did not understand very well the meaning of most of the anti-COVID-19 measures. This weakness needs to be addressed not only from the perspective of the population, but also from how such measures were defined by the government. For example the measure of keeping distance from each other was disseminated as “social distancing” first, but later it changed to “physical distancing”, and the minimum distance was between 1 and 1.5 meters before 2 meters came out (but the 2 meters was seldom disseminated). These changes occurred also in regard to “crowded places”-it was defined depending on whether the people are in a closed or open space, which was used to define the maximum number of people that could gather (including for funeral), and these numbers have been changing constantly. These unclear and/or constantly changing measures may have confused the population.

Compliance with anti-COVID-19 measures at individual and at household levels is low, both in Manhiça and in Quelimane – below 50% in most of the indicators used in this study – see percentage of “Yes” in table 7 and of those implementing a certain measure at 100% in table 6. A similar result (55.5% of overall level of compliance) was found in a study on the compliance with COVID-19 preventive measures among food and drink establishments in Ethiopia, in 2020 (25). Our study found that even the apparently easiest measure (mask wearing in public places), most people (50.8%) confessed that they did not comply with it when they think they should. Only 40.0% of the respondents said that they were avoiding unnecessary travels in 100% of the times, 34.0% were avoiding crowed places, and 29.1% were doing social distancing outside the household. Structural and personal factors may have contributed to lower compliance as most people live on daily-income activities and even those with a monthly income had to go for work using crowed public transports. This indicates that these measures could have been followed better by the population if the government had invested on public transports system to reduce overcrowding in the bus-stops and within the public transport vehicles.

The prevalence of symptoms of respiratory illness in the households was very low, and most respondents did not know anyone with such symptoms in their communities. However, the meaning or usefulness of these findings must be taken cautiously because of the bias involved in self-report of symptoms, when compared to medically diagnosed disease, particularly because some studies indicated that 80% of infected people in Mozambique were asymptomatic (13); six in seven COVID-19 infections went undetected in Africa (26), and that 75% of infected people in India were asymptomatic (27).

With regards to hardships imposed to people and their households by the pandemic, the study shows that the closure of educational institutions, fear of being infected, hunger, fear of death and of uncertainty of the future, and the closure of churches were the main hardships – and these responses seem expected, particularly for Manhiça and Quelimane where little or no social services exist for the most pre-COVID-9 vulnerable populations, whose suffering has increased due to Coronavirus. A study in Philippines on how COVID-19 impacted vulnerable communities, reported increased lack of income opportunities and insufficient food supply that existed before COVID-19 but had worsened due to the pandemic (28). One aspect deserving further research is why people are worried about the closure of churches and of *xitique*? Is it because they want to worship their God and receive the *xitique*-related savings or they want the entertainment or socialization that is involved in the churches and in *xitique*? These research questions may be important for mitigating COVID-19 impacts because the churches are institutions that mostly welcome suffering people and provide them with psychological and social strength that can improve the quality of their lives (29). Similarly, *xitique* provides the means for income savings and socialization not only for the least privileged, but also for relatively prosperous groups of individuals (20).

## 5. CONCLUSIONS

The awareness about Coronavirus was high both in Manhiça and in Quelimane districts (98.7% of participants had heard of Coronavirus), although the term COVID-19 was not as popular as that of Coronavirus (89.2%). Most participants knew about the disease and its transmission and prevention dynamics, but the level of understanding of anti-COVID-19 measures was low, as measured by the higher proportion of respondents that could not define the key measures accurately or fairly and of those who said that did not know how to define these measures. These low levels of perceptions suggest that the messages may not have been transmitted in an explanatory way enough for the general population to digest, particularly because some terms and rules were changing constantly over time, which may have confused the population.

The level of compliance with anti-COVID-19 measures was low (below 50% in most of the indicators used in this study) even for groups of individuals or of households that one would expect to find higher levels of compliance, such as urban population and households that have piped water. The measures that most people failed to comply with are social distancing within the household (77.4% of respondents said that they could not comply with it at all), followed by avoiding touching the month, nose, and eyes; staying at home, and frequent hand washing. The ratio of face masks per person aged 6 years or more is too low (1 mask per person), which suggests that people may be using the same mask for longer periods and/or repeatedly without replacing it, which is particularly worrying when considering that most people were using clothe masks whose efficacy is lower than medical masks. Structural and personal factors may have contributed for this lower compliance as most people live on daily-income activities and even those with a monthly income had to go for work using crowed public transports. This indicates that these measures could have been followed better by the population if the government had invested on public transports system to reduce overcrowding in the bus-stops and within the public transport system. There is a need to respond to these findings with provision of more detailed information which way increase the people’s awareness and adherence to the efforts to fight this pandemic.

## Data Availability

These data are not openly accessible in a web address, but they may be obtained through a formal request sent to Godifre Capinga, accompanied by a proposal that will be analysed by CISM’s internal scientific and ethical committees.

## DECLARATIONS

### Ethics approval and consent to participate

This study was reviewed and approved by the CISM’s Internal Scientific Committee (CCI) and CISM’s Internal Bioethics Committee (CIBS) (approval ref. CIBS-CISM/028/2020). Before starting the interview, each participant was informed about the study objectives and procedures and the voluntary nature of his/her participation through information read out from a participant’s information sheet. Participants were enrolled in the survey after obtaining their oral informed consent, which was voice-recorded (not paper-based consents) to minimize the risk of COVID-19 transmission associated with exchanging papers and pens or stamps for finger print between fieldworkers and participants.

### Consent for publication

The participant’s information sheet requested permission for publication of the data collected in a non-individualized manner.

### Authors’ contributions

AN: Participated in the study design and fieldwork, supported in the data cleaning and he conducted the analysis and interpretation of the data, and wrote this article;

AM: Participated in the study design and in the implementation of field activities and supervision of data collection, and contributed to the writing of this article;

FA: Participated in the study design and in the implementation of field activities and contributed to the writing of this article;

AH: Participated in the data management, including data processing and cleaning;

TM: Participated in the data management, including data processing and cleaning;

ArsN: Participated in the design of the study and supported the digitalization of the questionnaires and data cleaning, and contributed to the writing of this article;

QB: Contributed to the writing of this article;

CS: Contributed to the writing of this article;

IM: Contributed to the writing of this article;

KM: Conceived the study, participated in the study design interpretation of results and writing of this article;

All authors read and approved the final manuscript.

## Acknowledgements

The authors would like to thank the community and the community leaders, the members of the Community Advisory Board and the local authorities, including the District Health Authorities, the District Governments and the Municipal Authorities in Manhiça and Quelimane, and all our national and international partners for their dedication and collaboration.

